# Operationalizing the neural exposome for brain health and Alzheimer’s Disease and Related Dementias (AD/ADRD) vulnerability in rural settings: pilot study

**DOI:** 10.64898/2026.05.21.26353825

**Authors:** Juliana N Souza-Talarico, Hans-Joachim Lehmler, Jessica K Caldwell, Yamnia Cortes, Megan Zuelsdorff, Yuxin Fun, Jennie Embree, Cynthia Doyle, Karla Halverson, Maria Martinez Rangel, Alaa Harb, Olivia Croskey, Katherine Britt, Chelsea Howland, Ana Werneck Capuano

## Abstract

**INTRODUCTION:** Alzheimer’s disease and related dementias (AD/ADRD) arise from cumulative environmental, social, behavioral, and biological influences across the life course. The neural exposome framework conceptualizes how exogenous, behavioral, and endogenous factors interact to shape brain health; however, its application to preclinical AD/ADRD research, particularly in rural populations, remains limited.

**METHODS:** We developed and piloted a community-embedded, decentralized research model to operationalize the neural exposome framework among cognitively unimpaired adults aged 45+ in two rural Midwestern U.S. communities, integrating environmental, social, behavioral, geospatial, and biological measures to evaluate exposure-related neurobiological and cognitive vulnerability.

**RESULTS:** This approach demonstrated high feasibility and acceptability, achieving strong recruitment, retention, data completeness, and multidomain biomarker collection in rural community-based settings

**DISCUSSION:** Pilot findings support the feasibility of neural exposome-informed research in rural U.S. communities and highlight its potential to advance prevention-oriented research on brain health and AD/ADRD.

## 1. Background

An increasing body of evidence suggests that Alzheimer’s disease and related dementias (AD/ADRD) are shaped by modifiable factors across the life course, including environmental, contextual, and individual determinants that contribute to excess risk in underserved populations^1^. In response, prevention-oriented research has expanded, emphasizing individual-level behavioral interventions such as physical activity, diet, sleep, and cognitive engagement^2,3^. While important, these approaches do not fully account for the environmental, psychosocial, and structural contexts in which behaviors occur or the ways non-genetic vulnerability is shaped^4,5^, and their combined effects on brain health remain underrepresented in preclinical AD research.

The exposome, defined as the totality of environmental exposures across the life course, provides a framework for understanding how cumulative environmental, social, and behavioral influences interact with biological systems^5,6^. The neural exposome extends this concept to examine how exogenous, behavioral, and biological exposures influence brain health through neurobiological pathways and gene expression across the lifespan^7^. Given the plasticity of the nervous system, these exposures may accumulate and interact with genetic susceptibility to shape vulnerability to AD/ADRD^7^.

Recent initiatives, such as Expo AD and the Gateway Exposome Coordinating Center, have advanced exposome-informed research through data integration, geospatial characterization, and cross-study harmonization^4,8,9^. However, operationalizing neural exposome frameworks for AD/ADRD prevention remains challenging. Many studies focus on single exposure domains with limited integration across environmental, behavioral, and biological factors^10–12^. Exposures are also often examined independently of the neurobiological systems through which they influence cognitive aging, limiting mechanistic insight and early identification of vulnerability pathways, particularly during preclinical stages^4,13–16^.

The effects of cumulative exposures are shaped by geographic context, which influences environmental conditions, social structures, psychological stressors, and behavioral constraints^17^. Urban and rural environments differ substantially in these factors, challenging uniform prevention strategies^18^. However, geographic context remains insufficiently integrated into exposome-informed AD/ADRD research.

This gap is particularly evident in rural populations, where exposures may reflect agricultural practices, water contamination, healthcare access, geographic isolation, and socioeconomic disadvantage, alongside population aging and workforce shortages^18–21^. Despite these factors, rural populations remain underrepresented in biomarker-rich AD/ADRD research due to logistical and structural barriers, including geographic dispersion, limited transportation, restricted access to academic centers, and mistrust of research institutions^22–24^. These challenges are amplified in studies requiring biospecimen collection, repeated assessments, and extended visits.

The methodological demands of neural exposome research further complicate implementation. Understanding how cumulative exposures become biologically embedded requires integration of environmental, social, behavioral, and geospatial data with neurobiological measures across multiple levels. Such approaches are difficult to implement in geographically dispersed settings with limited research infrastructure. As a result, populations with context-specific exposure risks are often least accessible through traditional research models.

Addressing these challenges requires scalable, community-embedded research frameworks that expand access and enable multidomain data integration^25,26^. In this context, community engagement functions as a core methodological infrastructure rather than a supplementary recruitment strategy, supporting feasibility, trust, and sustained participation essential for longitudinal prevention research^25,27^.

The aims of this study were to (i) develop and describe a research infrastructure model to operationalize the neural exposome framework for brain health and AD/ADRD vulnerability in rural settings, and (ii) implement and evaluate this model in a pilot study to assess the feasibility and acceptability of community-based recruitment, decentralized multidomain exposure assessment, and biospecimen collection in non-clinical environments.

## 2. Method

### 2.1. Study Design

This study employed a developmental and implementation-oriented design to develop and pilot-test a research infrastructure model that operationalizes the neural exposome framework. The developmental phase focused on four components: (1) tailoring the neural exposome framework to address brain health and AD/ADRD vulnerability in rural settings; (2) specifying the research infrastructure required to support a neural exposome-informed study protocol; (3) designing a multidomain measurement architecture to integrate exogenous, behavioral, and endogenous exposures within rural contexts; and (4) defining indicators to evaluate feasibility and acceptability of both the research infrastructure and measurement approach. The implementation phase involved operationalizing this model in rural communities through research team training, establishing partnerships with community organizations, and deploying the study protocol. This phase was designed to evaluate feasibility and acceptability across key processes, including recruitment, enrollment, multidomain data completeness, and the operationalization of research infrastructure.

### 2.2 Developmental Phase

#### 2.2.1. Tailoring the Neural Exposome Framework for Rural AD/ADRD Contexts

The neural exposome framework conceptualizes brain health as the result of dynamic interactions among non-genetic exposures across the life course^7^. These exposures are organized into three interrelated domains, exogenous, behavioral, and endogenous, representing interconnected layers of influence on neural systems^7^. Exogenous exposures include environmental and contextual conditions, such as physical and chemical agents (e.g., pollutants, toxicants, light, noise) and broader structural factors, including climate, built environment, and socioeconomic context^7^.

Behavioral exposures encompass individual-level activities and experiences, including psychosocial stress, health behaviors (e.g., physical activity, sleep, diet), substance use, and social interactions, which translate external conditions into biological effects^7^. Endogenous exposures represent internal biological states and regulatory processes, including genetic and epigenetic mechanisms, metabolism, immune function, and hormonal signaling, through which cumulative exposures become biologically embedded and influence brain function and disease vulnerability^7^.

These domains operate as an integrated multilevel system characterized by continuous interaction and feedback^7^. Exogenous conditions shape behavior; behavioral processes modulate biological pathways; and endogenous systems influence how exposures are processed and expressed^7^.

Brain health and disease risk emerge from cumulative, time-dependent interactions across these levels rather than isolated factors^7^. Within this framework, neural vulnerability and plasticity influence how exposures are integrated biologically, positioning the central nervous system as an adaptive interface linking external conditions with internal regulatory systems across the lifespan^7^.

In the context of AD/ADRD vulnerability, this framework requires multidomain, multilevel assessment capturing how exposures co-occur, interact, and accumulate to influence neurobiological regulatory systems relevant to preclinical cognitive aging. This approach prioritizes identification of exposure patterns influencing biologically responsive pathways rather than estimating independent effects of single exposures^4,5,13^. Multidomain assessment is particularly informative prior to cognitive impairment, when biological signals more likely reflect environmentally driven dysregulation rather than downstream disease processes that alter behavior, physiology, and social functioning (e.g., reduced mobility, altered activity patterns, changes in stress physiology, and healthcare engagement)^15,16^. Once impairment develops, these changes can modify both exposure patterns and biological measures, introducing reverse causation and complicating causal inference. Focusing on cognitively unimpaired adults reduces such confounding and strengthens identification of exposure-responsive pathways relevant to early risk detection and prevention^1^.

Multidomain exposome research requires measurement across multiple levels of influence, including individual, interpersonal, and structural contexts. Individual-level measures capture internal dose and lived experience (e.g., chemical burden, perceived stress, sleep, physical activity), interpersonal measures reflect relational environments shaping stress physiology and resilience (e.g., social support), and community-level indicators provide essential context for interpreting exposure profiles, including healthcare access, environmental quality, occupational factors, and socioeconomic conditions^13^.

To capture these exposures, multidomain measurement extends beyond self-report and biospecimens to include passive and contextual data streams. Remote digital measures of behavior and physiology (e.g., physical activity, sleep, mobility) and geospatial indicators (e.g., air quality, land use, water systems) enable objective assessment of time-varying, cumulative, and co-occurring exposures within participants’ residential and activity spaces^5,13^. These data complement individual-level measures by capturing environmental and structural conditions not observable through questionnaires or clinic-based assessments.

In rural contexts, these data are particularly informative when integrated with individual experiential measures, as exposures may arise from agricultural activities, private water systems, occupational conditions, geographic isolation, limited access to healthcare and social services, and extended travel distances shaping daily routines and stress exposure^18,19^. Integrating these data streams supports objective characterization of place-based and structural influences and strengthens inference regarding interactions between environmental exposures and neuroendocrine and neuroimmune regulation during preclinical cognitive aging.

Neural exposome-informed measurement must also account for vulnerability and resilience. Psychosocial resources, including social support, sense of purpose, spirituality, and community connectedness, may moderate neurobiological responses to cumulative exposures and contribute to heterogeneity in cognitive trajectories^28,29^. Capturing these factors is essential for distinguishing adaptive from maladaptive responses and identifying targets for prevention.

Integrating multidomain and multilevel assessments during preclinical stages provides a mechanistically informed approach to understanding how real-world exposures and behaviors become biologically embedded across the life course, linking exogenous and behavioral factors to neurobiological regulation and early cognitive outcomes in prevention-oriented AD/ADRD research.

#### 2.2.2 Specifying Research Infrastructure for Neural Exposome Implementation

Within the neural exposome framework, feasibility and participant engagement are core elements of scientific infrastructure rather than ancillary operational considerations, as they are essential for valid assessment of multidomain, multilevel exposures across the life course. Neural exposome-informed AD/ADRD research requires coordinated integration of environmental, psychosocial, behavioral, biological, and cognitive data across heterogeneous real-world contexts4. When this integration cannot be consistently operationalized, exposure complexity is reduced, contextual heterogeneity is obscured, and relationships between lived environments and neurobiological regulation become difficult to interpret longitudinally.

Traditional clinic-based research models constrain such integration by centralizing study activities, which can fragment data collection and limit participation, particularly among rural and mobility-limited populations^22,23,30^. Geographic distance between communities and academic centers may also hinder sustained academic–community partnerships, contributing to communication gaps and reinforcing mistrust of research institutions^22,23^. In this context, feasibility is not solely a recruitment outcome but a determinant of whether multidomain exposome investigations can be conducted with sufficient depth and continuity.

To address these constraints, decentralized research methodologies, adapted from decentralized clinical trial (DCT) approaches, were applied to observational and longitudinal research in rural settings. These approaches redistribute study activities from centralized academic sites to participants’ homes or community settings using mobile, digital, and local healthcare resources, aiming to reduce access barriers while maintaining protocol standardization and data quality^31–33^. Data collection within this model is modular and participant-centered, combining community-based assessments with remote components tailored to participants’ characteristics, access, and preferences.

Decentralized infrastructures also support distributed longitudinal data acquisition by integrating geocoding, digital platforms, mobile devices, GIS linkages, and environmental databases^7,14^.

These capabilities are critical for neural exposome research, where geospatial and contextual data are required to capture environmental and structural exposures not observable within clinic-based designs. Within this infrastructure, diverse data collection strategies—including REDCap-based surveys^34^, digitally administered cognitive assessments^31–33^, and flexible biospecimen collection approaches—can be integrated into a unified system^35–37^.

Biospecimen strategies must balance participant feasibility with biomarker validity, as some AD/ADRD markers require specific collection and processing conditions not fully supported across decentralized modalities^36^. Accordingly, mobility is conceptualized not as a logistical convenience but as a methodological mechanism to preserve multidomain exposure characterization, improve representativeness, and strengthen ecological validity^27,38^.

However, expanded accessibility alone does not ensure meaningful engagement, institutional trust, or sustained participation, which are essential for longitudinal neural exposome research, particularly in underserved populations30,39. Therefore, decentralized infrastructure must be integrated with community-participatory research principles^40^, forming a community-embedded, decentralized research (CEDR) model. Community-embedded approaches incorporate core CBPR elements, including shared research priorities, recognition of community knowledge, and capacity building through structures such as community advisory boards (CABs). CEDR extends CBPR by decentralizing the location of data collection, embedding measurement activities within community settings rather than centralized academic or clinical environments.

While CBPR emphasizes equitable participation in guiding research, CEDR additionally shifts where data are generated, enhancing accessibility, ecological validity, and population reach. Within the CEDR model, community engagement functions as both a methodological strategy and an exposome-relevant process influencing feasibility, data quality, and interpretability. CABs include community organizations, middle-aged and older adults, healthcare providers, agricultural workers, and other local stakeholders, supporting identification of locally relevant exposures, risks, and resilience factors not captured by standardized measures^25,26^.

CABs also inform study design, measurement selection, and ethical governance, including biospecimen acceptability, longitudinal participation, and data stewardship expectations.

Incorporating these perspectives enhances feasibility while ensuring alignment between scientific constructs and lived experience^25,27^. Sustained investigator presence and relational continuity within communities through repeated engagement are similarly critical for building trust and supporting recruitment, retention, and long-term participation^26,39^.

Finally, bidirectional dissemination and education are integral components of feasibility. Returning aggregate findings, using study encounters to provide brain health education, and engaging communities in interpretation support sustained participation and iterative refinement of measurement strategies. Within the CEDR model, dissemination is not a post-study activity but part of an ongoing feedback process linking scientific discovery with community relevance and long-term impact^25,27^.

#### 2.2.3 Designing a Multidomain Measurement Architecture for Rural Exposures

Guided by the neural exposome framework, multidomain measures were selected to capture key exogenous, behavioral, and endogenous exposures across individual, interpersonal, and community levels. Selection was informed by relevance to rural contexts and to pathways implicated in brain health and AD/ADRD vulnerability, while balancing comprehensiveness with participant burden to ensure feasibility in community-based settings. This section outlines the theoretical and methodological basis for measurement selection rather than empirical characterization; specific measures and operationalization are summarized in Table 1. Consistent with the decentralized design, data collection combined REDCap-based surveys (remote or on-site), digital cognitive assessments, and biospecimen collection using community-based, portable, and participant-directed approaches.

**Table 1.** Multidomain Measurement Architecture and Operationalization.

| Domain | Component | Measures | Operationalization (Data Collection) |
| --- | --- | --- | --- |
| Exogenous<br>(Individual-level) | Environmental exposure history | Residential history; water source and use; occupational history; proximity to agricultural/industrial activities; local use of fish and produce | Self-report questionnaires administered via REDCap or paper-based surveys |
|  | Chemical exposure (internal dose) | Persistent organic pollutants (PCBs, PFAS) | Serum-based laboratory assays from venipuncture samples collected during community visits |
| (Community-level) | Environmental and structural context | Neighborhood stressors, infrastructure, and environmental exposures | Geocoding of residential addresses linked to publicly available datasets (ZIP code, census tract) |
|  | Water system exposures | Municipal drinking water system data | Linkage to administrative datasets (e.g., state environmental databases) |
|  | Socioeconomic and structural vulnerability | Area Deprivation Index (ADI); CDC Social Vulnerability Index (SVI) | Geospatial linkage to census-derived composite indices |
|  | Rurality and geographic isolation | Rural–Urban Commuting Area (RUCA) classifications | Geospatial classification based on residential location |
| Behavioral | Psychosocial stress and adversity | Perceived stress; life events; adverse childhood experiences; discrimination; loneliness; depression symptoms | Validated self-report questionnaires (REDCap or paper-based) |
|  | Health behaviors | Sleep, physical activity, diet, substance use | Validated self-report instruments |
|  | Reproductive health | Reproductive history; menopause symptoms (when applicable) | Self-report questionnaires |

|  | Resilience and psychosocial resources | Social support; spirituality; self-compassion; cognitive and social engagement | Validated self-report scales |
| --- | --- | --- | --- |
| Endogenous (Neuroendocrine) | HPA-axis regulation | Cortisol awakening response (CAR); DHEAS | Salivary sampling at awakening, 30 and 45 minutes post-awakening (self-collected with diaries); plasma DHEAS from blood samples |
| (Neuroimmune / Neurodegenerative) | Inflammatory and neurobiological markers | Cytokines; amyloid; tau; neurofilament light chain; synaptic and glial markers | Plasma-based proteomic assays |
| (Systemic health) | Cardiovascular and metabolic regulation | BMI; blood pressure; heart rate; insulin; glucose; HbA1c; lipid profile | In-person clinical assessment and laboratory assays |
| Cognitive outcomes | Global and domain-specific cognition | NIH Toolbox Cognition Battery (fluid and crystallized cognition) | Digital administration (English/Spanish) with automated scoring system |

##### Exogenous Factors of the Neural Exposome

Exogenous factors were conceptualized as upstream environmental and socio-structural conditions shaping exposure contexts over time. In rural settings, exposures often reflect complex interactions between environmental conditions, occupational practices, and resource availability, including agricultural activity, locally sourced food systems, and reliance on private or small municipal water systems. These pathways contribute to cumulative exposures relevant to AD/ADRD risk but are often underrepresented in population-based research.

Measurement selection therefore integrated individual-level exposure data with community-level environmental context. Individual measures capture participant-specific exposures, while geospatial indicators provide contextualized information on environmental and structural conditions, including socioeconomic disadvantage, infrastructure, and geographic isolation (Table 1). These indicators function as upstream determinants shaping exposure opportunities and constraints, influencing behavioral processes and downstream biological regulation.

Integrating individual and place-based measures enables comprehensive characterization of how environmental and structural contexts interact with biologically responsive pathways during preclinical cognitive aging. This approach addresses a key gap in AD/ADRD research by incorporating place-based exposure patterns not captured in clinic-based studies.

##### Behavioral Factors of the Neural Exposome

Behavioral factors were conceptualized as processes through which external conditions are internalized and translated into physiological responses. These include psychosocial stressors, health behaviors, and social experiences shaping stress regulation, coping, and resilience (Table 1).

Measurement selection prioritized both burden-related and resilience-related constructs. Stress-related domains include chronic and acute stressors, adverse experiences, discrimination, and social isolation, while resilience domains include social support, engagement, spirituality, and adaptive psychological resources. These dimensions capture variability in how individuals respond to similar environmental conditions.

Behavioral processes function as intermediaries linking exposures to neurobiological regulation. Including both vulnerability and resilience pathways supports examination of how psychosocial and lifestyle factors modulate biological embedding of exposures relevant to cognitive aging and AD/ADRD risk.

##### Endogenous Factors of the Neural Exposome

Endogenous factors represent internal regulatory systems through which exposures are biologically embedded and expressed in pathways relevant to brain health and AD/ADRD vulnerability. This reflects the understanding that exposure effects are distributed across interacting systems rather than localized to a single target^7^.

Measurement selection prioritized neuroendocrine and neuroimmune systems as key interfaces linking exposures to downstream neural and cognitive outcomes (Table 1). These systems were selected due to their responsiveness to environmental and psychosocial exposures, their role in maintaining systemic and neural homeostasis, and their established relevance to cognitive aging and AD/ADRD vulnerability^39,41–56^.

Neuroendocrine processes, particularly glucocorticoid regulation, function as upstream pathways through which exposures such as chronic stress, structural adversity, and endocrine-disrupting chemicals influence neural function^42–45^. These pathways regulate metabolic, vascular, and synaptic processes and have been associated with alterations in cortico-limbic structures and cognitive decline in preclinical AD/ADRD^42–45^.

Neuroimmune processes represent the broader biological context in which exposures are expressed, reflecting states of vulnerability or resilience through inflammatory and glial-mediated pathways (Table 1). These systems integrate environmental, psychosocial, and metabolic influences and are implicated in early AD/ADRD mechanisms, including microglial activation and interactions with amyloid and tau pathology^39,46–48,41,49–56^.

Additional metabolic and cardiovascular measures were included to capture systemic processes that co-occur with and interact with neurobiological pathways (Table 1), providing a broader context for how cumulative exposures are expressed across physiological systems relevant to cognitive aging.

##### Cognitive Outcomes

Cognitive outcomes were conceptualized as downstream functional indicators reflecting cumulative effects of exogenous, behavioral, and endogenous processes. Rather than diagnostic endpoints, cognition is treated as an integrative outcome of exposure-related variability.

Measurement selection prioritized domains sensitive to early change, particularly those linked to stress-related biomarkers and preclinical neurodegenerative processes (Table 1). The NIH Toolbox Cognition Battery was selected based on its suitability for community-based research, strong psychometric properties, and ability to assess both fluid and crystallized cognition^57,58^.

Fluid cognition measures are sensitive to neuroendocrine, immune, and metabolic processes associated with early-stage AD/ADRD and exposure-related variability^59–64^. Crystallized cognition provides a stable reference reflecting accumulated knowledge and cognitive reserve, supporting interpretation of exposure-related differences^57^. Evidence linking NIH Toolbox measures to preclinical AD biomarkers further supports their relevance for prevention-oriented research^61–64^. Together, these measures situate cognitive function within a multidomain framework that captures the biological embedding of exposures and supports the evaluation of early variability in brain health related to AD/ADRD risk.

#### 2.2.4 Defining Feasibility and Acceptability Assessment

Feasibility and acceptability of the CEDR model were evaluated using a structured, multidomain approach combining quantitative implementation indicators and qualitative feedback. Feasibility was assessed across four domains: (1) recruitment reach and pathway distribution, (2) enrollment and study completion, (3) data completeness and protocol adherence, and (4) operational implementation of decentralized data collection. Acceptability was assessed using behavioral participation indicators and qualitative feedback.

Measurement procedures included REDCap-based data capture, participant questionnaires, sampling diaries, recruitment and screening logs, and study documentation guided by the manual of operations. Table 2 summarizes the domains, indicators, and data sources.

**Table 2.** Feasibility and Acceptability Domains, Indicators, and Measurement Procedures.

| **Domain** | **Indicators** | **Measurement Procedures** |
| --- | --- | --- |
| Recruitment reach and pathway distribution | Number of individuals reached; distribution across recruitment channels; number of neighborhoods within the county where data collection was conducted | Recruitment logs; REDCap screening records; participant self-report of recruitment source |
| Representation of target population | Rural residency; demographic characteristics (age, education, race/ethnicity); comparison with Census data | Participant questionnaires; geocoding (ZIP code, RUCA); Census data linkage |
| Enrollment and study completion | Progression through study stages; completion rates; retention; time to complete baseline procedures | REDCap enrollment tracking; study logs; visit scheduling records |
| Participant burden | Duration of study visits; completion of study tasks | Visit logs; study protocol tracking |
| Data completeness and protocol adherence | Proportion of complete datasets; missing data frequency; adherence to biospecimen protocols (e.g., CAR sampling) | REDCap real-time completion flags; follow-up logs; saliva collection diaries |
| Biospecimen feasibility | Sample completion; sample usability; absence of processing failures; biomarker detectability; processing time | Laboratory records; field collection logs; biospecimen processing logs; assay outputs |
| Acceptability (behavioral) | Completion of high-burden procedures; willingness for longitudinal follow-up; participation in digital assessments | REDCap surveys; completion logs; study records |
| Acceptability (qualitative) | Participant and community feedback on procedures, communication, and relevance | Informal field notes; staff observations; community partner feedback |
| Engagement and sustainability | Participation in dissemination activities; continued community interaction | Event attendance records; partnership communication logs |

Data collection was supported by integrated systems, including REDCap for surveys and enrollment tracking, structured study logs, participant-completed biospecimen diaries, and laboratory records. Real-time monitoring enabled identification of missing data and targeted follow-up, with procedures documented in study logs. Biospecimen adherence, including multi-timepoint cortisol sampling, was verified using time-stamped diaries and staff confirmation. Qualitative indicators of acceptability were captured through field notes and participant and community partner interactions.

These measures provide preliminary evidence of the model’s capacity to support recruitment, multidomain data collection, biospecimen acquisition, and participant engagement required to operationalize the neural exposome framework in rural AD/ADRD research. Consistent with a pilot design, metrics were descriptive and not intended to assess comparative effectiveness or cost-efficiency.

### 2.3 Implementation Phase

To assess the feasibility of applying the neural exposome framework in real-world rural settings, a pilot study was conducted using the CEDR infrastructure. Study procedures followed a structured sequence including (1) research team training, (2) community engagement, and (3) study protocol deployment, encompassing distributed recruitment, remote and in-person screening, and modular data collection.

#### 2.3.1 Research Team Composition and Training

Study procedures were implemented by a multidisciplinary team including licensed nurses, medical assistants, a doctoral-prepared nurse practitioner, and bachelor’s- and doctoral-level staff. Team members were selected based on clinical experience, research training, and community engagement expertise, with bilingual capacity in English and Spanish for participant-facing procedures. Additional language capacity (Portuguese, Chinese, Arabic) supported broader communication needs.

All team members completed standardized training covering study protocol, human subjects’ protections, cognitive assessment, biospecimen collection and handling (including transport and storage), bloodborne pathogen certification, cash handling, and community-engaged research practices. Training was delivered using in-person, synchronous, and asynchronous formats, including live virtual sessions and recorded materials. Instruction combined didactic sessions, review of the manual of operations, procedural demonstrations, and supervised field practice to ensure consistency across sites. Protocol fidelity was reinforced through team meetings, field supervision, and investigator oversight. Real-time monitoring using REDCap-based checklists enabled identification of missing procedures and timely resolution, promoting standardized data collection across decentralized settings.

#### 2.3.2 Establishing Community Partnerships

Community partnerships were central to the CEDR model, serving as operational anchors and governance structures. Collaborations with libraries, healthcare clinics, recreation centers, and nonprofit organizations supported study planning and implementation. Partners provided guidance on outreach, communication channels, site logistics, and dissemination, ensuring alignment with local context.

Stakeholders contributed to recruitment materials, language, and messaging prior to study launch to enhance cultural and contextual relevance. Educational materials were co-developed with a community advisory board (CAB) including healthcare providers and middle-aged and older adults from underserved and Hispanic communities. All materials were available in English and Spanish. Governance emphasized bidirectional communication and shared stewardship. Community partners were involved in protocol refinement, implementation, and problem-solving, allowing adaptation while maintaining scientific fidelity. Dissemination included workshops presenting aggregate findings to participating communities, supporting transparency and engagement.

Trust-building was treated as an active process. Study activities were conducted by a consistent team working with community facilitators, with multilingual capacity to support participants. Procedures were explained clearly, and participants were encouraged to ask questions to support autonomy and comprehension.

Sustained investigator presence further supported engagement. Investigators and coordinators maintained visibility through repeated visits and direct interaction, reinforcing study purpose and procedural consistency. Study procedures were designed to align with rural contexts through flexible scheduling, streamlined workflows, and sensitivity to research familiarity levels, supporting ongoing engagement beyond initial enrollment.

#### 2.3.3 Deploying the Study Protocol: Recruitment, Screening, and Data Collection

The study was conducted across two rural counties in a U.S. Midwest State selected based on scientific relevance and operational feasibility, including rural classification, demographic composition, and socioeconomic variation. Proximity to municipal water systems with documented environmental contaminants supported integration of geospatial and biological exposure data. Study sites were located within approximately 70 miles of the processing center to maintain biospecimen integrity.

Recruitment was conducted through community partnerships and outreach across healthcare settings, local businesses, public services, media, and social platforms, with additional engagement through organizations serving Hispanic/Latino communities. Study staff participated in community events to enhance visibility and engagement.

Eligibility screening was completed remotely via REDCap or through direct contact with study staff. This approach reduced burden and minimized unnecessary visits. Eligible participants completed cognitive screening and informed consent electronically or on paper based on preference.

Participants completed self-administered questionnaires via REDCap, with paper options available. Instructions for study procedures were provided verbally, in writing, and through video materials.

Communication and reminders were tailored to participant preference.

Data collection followed a modular, decentralized approach. In-person assessments were conducted through mobile visits at community sites such as libraries and municipal buildings. Study activities included cognitive testing, portable phlebotomy, and digital assessments.

Biospecimen collection used multiple approaches, including community-based, field-based, and home-based collection.

Participants received individualized reports summarizing clinical and behavioral measures, including lipid profiles, glucose levels, BMI, blood pressure, and cognitive screening results.

Reports included explanations and recommendations for follow-up when needed, including referral to healthcare providers or low-cost clinics. Brain health education was provided through verbal and printed materials.

Participants were compensated upon completion of study procedures in accordance with institutional review board approval (IRB# 202412177).

## 3. Results

### 3.1 Feasibility and Acceptability Outcomes

Feasibility and acceptability of the CEDR model were evaluated across recruitment, representation of the target population, enrollment and study completion, data completeness and protocol adherence, biospecimen feasibility, and participant engagement.

#### 3.1.2 Recruitment Reach and Representation

A total of 67 individuals were reached across seven rural communities in two counties, of whom 50 met eligibility criteria and were enrolled (conversion rate: 74.6%). Recruitment occurred through multiple community-based pathways, including interpersonal networks (31.9%), community events (29.0%), and flyers or posters (26.1%), with additional contributions from social media, healthcare settings, and local organizations, indicating distributed engagement across access points.

All enrolled participants resided in rural areas, confirmed through ZIP code linkage with U.S. Census and Rural–Urban Commuting Area (RUCA) classifications. The sample had a mean age of 63.9 ± 9.37 years (range 45.0–76.0), mean education of 13.0 ± 3.71 years (range 2.0–20.0), and mean length of residence of 20.9 ± 11.5 years (range 3.31–50.31), reflecting a mid- to older-age population with long-term community residence. Sociodemographic characteristics indicated diversity in race and ethnicity, with a predominantly White sample and substantial Hispanic/Latino representation. Socioeconomic indicators varied across income, household composition, and health insurance coverage, reflecting diverse living conditions and access to care. Household structure was largely composed of smaller households, consistent with later life stages.

Environmental indicators further reflected rural exposure contexts, with most participants residing near agricultural environments and a subset reporting agricultural work exposure (Table 3).

**Table 3.** Participant Sociodemographic and Socioeconomic Characteristics.

| **Variable** | **Total Sample** | **n** | **%** |
| --- | --- | --- | --- |
| **Race** |  |  |  |
| White | 43 | 31 | 72.1 |
| Black/African American | 43 | 1 | 2.3 |
| Asian | 43 | 1 | 2.3 |
| Mixed race | 43 | 4 | 9.3 |
| Prefer not to answer | 43 | 6 | 14.0 |
| **Ethnicity** |  |  |  |
| Hispanic/Latino | 46 | 27 | 58.7 |
| Non-Hispanic/Latino | 46 | 16 | 34.8 |
| **Household income** |  |  |  |
| ≥ \$80,000 | 47 | 19 | 40.4 |
| \$20,000–\$39,999 | 47 | 8 | 17.0 |
| < \$20,000 | 47 | 8 | 17.0 |
| \$40,000–\$79,999 | 47 | 7 | 14.9 |
| Prefer not to disclose | 47 | 5 | 10.6 |
| **Household size** |  |  |  |
| 1–2 persons | 47 | 30 | 63.8 |
| 3 persons | 47 | 9 | 19.1 |
| ≥ 4 persons | 47 | 8 | 17.1 |
| **Health insurance** |  |  |  |
| Full coverage | 48 | 36 | 75.0 |
| Limited coverage | 48 | 6 | 12.5 |
| No insurance | 48 | 5 | 10.4 |
| Prefer not to answer | 48 | 1 | 2.1 |
| Living near agricultural fields (yes) | 48 | 38 | 79.2 |
| Household exposure to agricultural employment (yes) | 48 | 14 | 29.2 |

Together, these findings demonstrate the capacity of the CEDR infrastructure to recruit and characterize a socioeconomically and demographically diverse rural population with varied environmental exposures and healthcare access contexts, providing preliminary evidence supporting application of the neural exposome framework in real-world rural settings (Table 3).

#### 3.1.3 Enrollment, Study Completion, and Participant Burden

Enrollment and participation feasibility were high. Of the 50 enrolled participants, 49 (98%) completed the full multidomain protocol; one participant did not complete biospecimen and cognitive assessments despite follow-up. Recruitment and enrollment were completed within 45 days, indicating efficient implementation of the decentralized model.

Procedures were completed within a coordinated timeframe, with a 3–7 day interval between screening and in-person assessments, reflecting operational efficiency and manageable participant burden. Screening and onboarding required approximately 30 minutes, and in-person visits approximately 75 minutes. High completion rates across procedures indicate acceptability in terms of time and complexity.

#### 3.1.4 Data Completeness and Protocol Adherence

Data completeness across survey and biological measures was high. Missing data were limited (n = 7) and primarily due to “prefer not to answer” responses for selected sociodemographic and economic items rather than incomplete participation. Real-time monitoring through REDCap enabled identification of missing data and targeted follow-up, resulting in near-complete datasets. Protocol adherence for biospecimen collection was also high. All participants completed multi-timepoint saliva sampling for the cortisol awakening response (CAR), with full compliance with sampling schedules and time-stamped documentation, demonstrating feasibility of temporally structured biospecimen protocols in decentralized settings.

#### 3.1.5 Biospecimen Feasibility

Biospecimen collection and processing were successfully implemented. All saliva and blood samples yielded valid assay results, with no loss, contamination, or processing failures. Samples were processed within approximately 60 minutes, consistent with assay requirements. Biomarker detectability was high (∼95%), with rates ranging from 85–100% for persistent organic pollutants (PFAS), supporting sample integrity under community-based conditions.

#### 3.1.6 Acceptability and Engagement

Acceptability of the CEDR model was supported by high participation and engagement. All participants (100%) expressed willingness to participate in future longitudinal assessments, indicating strong acceptability of study procedures, including fasting biospecimen collection, multidomain questionnaires, and digital cognitive testing.

Qualitative feedback indicated that participants perceived study procedures as relevant, understandable, and acceptable. Digital cognitive assessments were positively received, with participants noting reduced perceived burden compared with examiner-administered approaches while maintaining adequate support from research staff.

## 4. Discussion

In this pilot implementation across two rural Midwestern counties, the CEDR model provided preliminary evidence of feasibility for operationalizing a multidomain neural exposome framework in community settings. The model supported geographically distributed recruitment, high retention and protocol adherence, and implementation of multidomain data collection spanning exogenous, behavioral, and biological exposures. These findings suggest that community-embedded, decentralized approaches may be a viable strategy for neural exposome–informed AD/ADRD research in rural populations.

CEDR enabled recruitment through multiple community-based pathways, facilitating access beyond centralized clinical systems. The sample reflected key sociodemographic and environmental characteristics of rural Midwestern populations, including long-term residential stability, heterogeneous income distribution, variability in healthcare coverage, and substantial proximity to agricultural environments. These features suggest the approach was effective in reaching populations underrepresented in biomarker-intensive aging research^21,24,27^.

Across feasibility domains, retention and completion of multidomain assessments were high, supporting implementation of complex protocols in community settings without substantial loss of data completeness. These findings should be interpreted as preliminary, given the pilot design.

High adherence to temporally structured biospecimen protocols, including neuroendocrine sampling, and successful processing indicate that rigorous biological data collection can be achieved outside clinical environments when supported by standardized protocols and real-time monitoring systems^31–33^.

Integration of environmental, socioeconomic, behavioral, and biological measures further supports feasibility of applying the neural exposome framework in real-world contexts. Observed variability across domains underscores the importance of multidomain exposure assessment, particularly for rural populations where environmental and structural exposures are often underrepresented in clinic-based cohorts^21,24,27^. However, these observations remain descriptive given the pilot scope. Cognitive testing using the NIH Toolbox digital platform, supported by on-site staff, was well accepted, with participant feedback indicating accessibility and engagement consistent with prior reports^61–62^. High willingness to participate in future waves suggests strong acceptability, although longitudinal retention remains to be evaluated. Together, these findings provide preliminary support for the potential scalability of community-embedded decentralized approaches for AD/ADRD prevention research in rural settings.

### 4.1 Lessons Learned

A key observation is that participant willingness was not a limiting factor for multidomain, biomarker-rich research. High enrollment and near-complete data acquisition indicate strong acceptability among middle-aged and older adults in participating rural communities. Feasibility was further supported by efficient operational processes, including streamlined scheduling, coordinated procedures, and minimization of logistical barriers. These findings challenge assumptions that rural populations are difficult to engage in complex biomedical research and instead highlight the importance of study design and implementation capacity^25,27,30^.

Recruitment patterns indicated that participation was strongly influenced by social networks and community interaction rather than institutional referral systems, suggesting an important role for trust and relational diffusion in recruitment^25,27,30^. A multidisciplinary team, including clinicians, behavioral scientists, implementation scientists, and community-engaged researchers, supported this process. Multilingual staff and community connections likely enhanced accessibility and trust. Multidisciplinary advisory structures and community partnerships strengthened implementation, including measure selection, quality control, and recruitment logistics. Community stakeholders contributed to culturally appropriate recruitment and dissemination strategies, consistent with CBPR principles^25–26^.

Sustained investigator presence further supported engagement and transparency, reinforcing study purpose and fostering trust. While these observations suggest the importance of relational continuity, further research is needed to quantify its impact on retention and data quality.

High data completeness was supported by real-time REDCap monitoring, structured follow-up, and standardized procedures, highlighting the importance of data infrastructure and training in decentralized research^33–34^.

Integration of individual-level data with geospatial and structural indicators improved contextual interpretation of exposures, particularly where self-reported socioeconomic data were incomplete. However, further methodological work is needed to validate approaches for integrating multilevel data.

### 4.3. Limitations

Several limitations should be considered. First, the pilot design, small sample size, and restriction to two rural counties limit generalizability. Second, although overall completeness was high, socioeconomic variables (e.g., race/ethnicity, income) had higher missingness, consistent with known challenges in self-reported sensitive data^67^, supporting the value of complementary geospatial indicators. Third, the feasibility assessment focused on recruitment, retention, and data completeness and did not include a formal evaluation of cost-effectiveness, scalability, or long-term sustainability. Fourth, longitudinal retention across repeated waves was not assessed.

Finally, although multidomain data collection was achieved, further work is needed to develop analytic approaches for integrating exposome data across domains.

### 4.4 Implications for Future AD/ADRD Research

These findings suggest that multidomain exposome assessment can be extended beyond clinic-based cohorts to rural populations using community-embedded, decentralized infrastructure. This approach enables better characterization of environmental, behavioral, and biological exposures in real-world contexts^4,7^.

Future research should evaluate scalability, sustainability of longitudinal implementation, and integration with existing cohort infrastructures. Advancing exposome research will also require analytic frameworks capable of integrating multidomain data, including modeling of complex interactions, harmonization across data types, and development of visualization tools. Integration of passive monitoring and digital health approaches may further enhance exposure characterization over time^31^.

Embedding research within community contexts remains essential not only for recruitment and retention but also for ensuring that exposure assessment reflects lived environments and promotes equity in AD/ADRD prevention research.

## 5. Conclusion

Advancing AD/ADRD prevention research requires both improved characterization of environmental and biological risk factors and research infrastructures capable of capturing these exposures in real-world settings. This pilot provides preliminary evidence that a community-embedded, decentralized approach can operationalize the neural exposome framework in rural populations while maintaining high data completeness, engagement, and methodological rigor. By aligning multidomain frameworks with pragmatic implementation strategies, the CEDR model offers a promising foundation for future longitudinal and intervention studies addressing exposure-related risk for cognitive decline in underserved populations.

## Data Availability

De-identified data that support the findings of this study are available from the corresponding author upon reasonable request, subject to institutional approval and data use agreements to ensure compliance with institutional policies and participant privacy protections

## ACKNOWLEDGMENTS

The authors gratefully acknowledge the community organizations that partnered in this research and provided trusted, accessible spaces for recruitment and data collection, including local public libraries, a university-affiliated primary care clinic, and the Latino community-based organization. We also extend our sincere appreciation to the community members who participated in the study and generously contributed their time and trust to this research. We thank the University of Iowa Environmental Health Research Center for methodological and scientific support.

## CONFLICTS OF INTEREST

The authors have no conflicts of interest to disclose.

## FUNDING SOURCES

Research reported in this publication was supported by the Csomay Center Optimal Aging pilot funding and National Center for Advancing Translational Sciences of the National Institutes of Health under Award Number UM1TR004403. The content is solely the responsibility of the authors and does not necessarily represent the official views of the National Institutes of Health.

## CONSENT STATEMENT

Participants provided written informed consent prior to enrollment in the study

## Notes

### Competing Interest Statement

The authors have declared no competing interest.

### Author Declarations

University of Iowa IRB gave ethical approval for this work (IRB# 202412177)

## References

1. Livingston G, Huntley J, Liu KY, et al. Dementia prevention, intervention, and care: 2024 report of the Lancet Standing Commission. Lancet. 2024;404(10452):572–628. doi:10.1016/S0140-6736(24)01296-0

2. Baker LD, Espeland MA, Whitmer RA, et al. Structured vs self-guided multidomain lifestyle interventions for global cognitive function: the US POINTER randomized clinical trial. JAMA. 2025;334(8):681–691. doi:10.1001/jama.2025.12345

3. Ngandu T, Lehtisalo J, Solomon A, et al. A 2-year multidomain intervention of diet, exercise, cognitive training, and vascular risk monitoring versus control to prevent cognitive decline in at-risk elderly people (FINGER): a randomised controlled trial. Lancet. 2015;385(9984):2255–2263. doi:10.1016/S0140-6736(15)60461-5

4. Finch CE, Kulminski AM. The Alzheimer’s disease exposome. Alzheimers Dement. 2019;15(9):1123–1132. doi:10.1016/j.jalz.2019.06.3910

5. Wild CP. The exposome: from concept to utility. Int J Epidemiol. 2012;41(1):24–32. doi:10.1093/ije/dyr236

6. Wild CP. Complementing the genome with an “exposome”: the outstanding challenge of environmental exposure measurement in molecular epidemiology. Cancer Epidemiol Biomarkers Prev. 2005;14(8):1847–1850. doi:10.1158/1055-9965.EPI-05-0456

7. Tamiz AP, Koroshetz WJ, Dhruv NT, Jett DA. A focus on the neural exposome. Neuron. 2022;110(8):1286–1289. doi:10.1016/j.neuron.2022.03.013

8. Cheng Y, Lyons D, Thursday N, DeWitt A, Bendlin BB, Kind AJ. Expo-AD: an innovative Alzheimer’s disease and related dementias (ADRD) research infrastructure for secure data integration and exposome linkage. Alzheimers Dement. 2024;20:e095640. doi:10.1002/alz.095640

9. Gateway Exposome Coordinating Center. Approach. Gateway Exposome Coordinating Center. Accessed May 17, 2026. https://gatewayexposome.org/#approach

10. Legaz A, Moguilner S, Barttfeld P, et al. The exposome of brain aging across 34 countries. Nat Med. 2026;32:1–14. doi:10.1038/s41591-026-00000-0

11. Weuve J, Puett RC, Schwartz J, Yanosky JD, Laden F, Grodstein F. Exposure to particulate air pollution and cognitive decline in older women. Arch Intern Med. 2012;172(3):219–227. doi:10.1001/archinternmed.2011.683

12. Hernandez H, Santamaria-Garcia H, Moguilner S, et al. The exposome of healthy and accelerated aging across 40 countries. Nat Med. 2025;31(9):3089–3100. doi:10.1038/s41591-025-00000-0

13. Vermeulen R, Schymanski EL, Barabási AL, Miller GW. The exposome and health: where chemistry meets biology. Science. 2020;367(6476):392–396. doi:10.1126/science.aay3164

14. Ibáñez A, Duran-Aniotz C, Migeot J, et al. Computational whole-body-exposome models for global precision brain health. Nat Commun. 2025;16(1):11078. doi:10.1038/s41467-025-00000-0

15. Bateman RJ, Xiong C, Benzinger TLS, et al. Clinical and biomarker changes in dominantly inherited Alzheimer’s disease. N Engl J Med. 2012;367(9):795–804. doi:10.1056/NEJMoa1202753

16. Jack CR Jr, Bennett DA, Blennow K, et al. NIA-AA research framework: toward a biological definition of Alzheimer’s disease. Alzheimers Dement. 2018;14(4):535–562. doi:10.1016/j.jalz.2018.02.018

17. Stingone JA, Geller AM, Hood DB, et al. Community-level exposomics: a population-centered approach to address public health concerns. Exposome. 2023;3(1):osad009. doi:10.1093/exposome/osad009

18. Kramer M, Cutty M, Knox S, Alekseyenko AV, Mollalo A. Rural-urban disparities of Alzheimer’s disease and related dementias: a scoping review. Alzheimers Dement (N Y). 2025;11(1):e70047. doi:10.1002/trc2.70047

19. Mollalo A, Kramer M, Cutty M, Hoseini B. Systematic review and meta-analysis of rural-urban disparities in Alzheimer’s disease dementia prevalence. J Prev Alzheimers Dis. 2025:100305. doi:10.14283/jpad.2025.100305

20. Morales DA, Barksdale CL, Beckel-Mitchener AC. A call to action to address rural mental health disparities. J Clin Transl Sci. 2020;4(5):463–467. doi:10.1017/cts.2020.42

21. Rahman M, White EM, Thomas KS, Jutkowitz E. Assessment of rural-urban differences in health care use and survival among Medicare beneficiaries with Alzheimer disease and related dementia. JAMA Netw Open. 2020;3(10):e2022111. doi:10.1001/jamanetworkopen.2020.22111

22. Friedman DB, Foster C, Bergeron CD, Tanner A, Kim SH. A qualitative study of recruitment barriers, motivators, and community-based strategies for increasing clinical trials participation among rural and urban populations. Am J Health Promot. 2015;29(5):332–338. doi:10.4278/ajhp.130514-QUAL-247

23. Kim SH, Tanner A, Friedman DB, Foster C, Bergeron CD. Barriers to clinical trial participation: a comparison of rural and urban communities in South Carolina. J Community Health. 2014;39(3):562–571. doi:10.1007/s10900-013-9798-2

24. Hoffman R, Choquette E, Blocker E, Murphy B, Howren B, Vidoni E. Rural health disparities in Alzheimer’s disease and related dementias biomarker testing. Innov Aging. 2025;9(suppl 2):igaf122.024. doi:10.1093/geroni/igaf122.024

25. Wallerstein N, Duran B, Oetzel JG, Minkler M. Community-Based Participatory Research for Health: Advancing Social and Health Equity. 3rd ed. John Wiley & Sons; 2017.

26. Israel BA, Coombe CM, Cheezum RR, et al. Community-based participatory research: a capacity-building approach for policy advocacy aimed at eliminating health disparities. Am J Public Health. 2010;100(11):2094–2102. doi:10.2105/AJPH.2009.170506

27. Parker EA, Baquero B, Daniel-Ulloa J, et al. Establishing a community-based participatory research partnership in a rural community in the Midwest. Prog Community Health Partnersh. 2019;13(2):201–208. doi:10.1353/cpr.2019.0019

28. Norton S, Matthews FE, Barnes DE, Yaffe K, Brayne C. Potential for primary prevention of Alzheimer’s disease: an analysis of population-based data. Lancet Neurol. 2014;13(8):788–794. doi:10.1016/S1474-4422(14)70136-X

29. Lee M, Whitsel E, Avery C, et al. Variation in population attributable fraction of dementia associated with potentially modifiable risk factors by race and ethnicity in the US. JAMA Netw Open. 2022;5(7):e2219672. doi:10.1001/jamanetworkopen.2022.19672

30. Bonevski B, Randell M, Paul C, et al. Reaching the hard-to-reach: a systematic review of strategies for improving health and medical research with socially disadvantaged groups. BMC Med Res Methodol. 2014;14(1):42. doi:10.1186/1471-2288-14-42

31. Cummins MR, Burr J, Young L, et al. Decentralized research technology use in multicenter clinical research studies based at US academic research centers. J Clin Transl Sci. 2023;7(1):e250. doi:10.1017/cts.2023.674

32. Dulko D, Kwong M, Palm ME, Trinquart L, Selker HP. From a decentralized clinical trial to a decentralized and clinical-trial-in-a-box platform: towards patient-centric and equitable trials. J Clin Transl Sci. 2023;7(1):e236. doi:10.1017/cts.2023.660

33. Apostolaros M, Babaian D, Corneli A, et al. Legal, regulatory, and practical issues to consider when adopting decentralized clinical trials: recommendations from the Clinical Trials Transformation Initiative. Ther Innov Regul Sci. 2020;54(4):779–787. doi:10.1007/s43441-019-00027-y

34. Harris PA, Taylor R, Thielke R, Payne J, Gonzalez N, Conde JG. Research electronic data capture (REDCap)-a metadata-driven methodology and workflow process for providing translational research informatics support. J Biomed Inform. 2009;42(2):377–381. doi:10.1016/j.jbi.2008.08.010

35. McDade TW, Williams S, Snodgrass JJ. What a drop can do: dried blood spots as a minimally invasive method for integrating biomarkers into population-based research. Demography. 2007;44(4):899–925. doi:10.1353/dem.2007.0038

36. Schultz AA, Paulsen AJ, Fredricks A, Plante DT, Peppard PE, Wilson R. Feasibility and validity of using self-collected capillary blood using Tasso+ for measuring Alzheimer’s disease plasma-based biomarkers among underrepresented populations. medRxiv. Published online February 2, 2026. doi:10.1101/2026.02.02.26345372

37. Emami A, Engström G, Kim H. Feasibility of in-home saliva collection of cortisol and DHEA-S as a biomarker of stress in dementia care dyads. Innov Aging. 2021;5(suppl 1):1005. doi:10.1093/geroni/igab046.3676

38. Islam NS, Zanowiak JM, Wyatt LC, et al. A randomized-controlled, pilot intervention on diabetes prevention and healthy lifestyles in the New York City Korean community. J Community Health. 2013;38(6):1030–1041. doi:10.1007/s10900-013-9710-0

39. Slavich GM, Irwin MR. From stress to inflammation and major depressive disorder: a social signal transduction theory of depression. Psychol Bull. 2014;140(3):774–815. doi:10.1037/a0035302

40. Kwon SC, Tandon SD, Islam N, Riley L, Trinh-Shevrin C. Applying a community-based participatory research framework to patient and family engagement in the development of patient-centered outcomes research and practice. Transl Behav Med. 2018;8(5):683–691. doi:10.1093/tbm/ibx026

41. Oberheim NA, Takano T, Han X, et al. Uniquely hominid features of adult human astrocytes. J Neurosci. 2009;29(10):3276–3287. doi:10.1523/JNEUROSCI.4707-08.2009

42. Lupien SJ, McEwen BS, Gunnar MR, Heim C. Effects of stress throughout the lifespan on the brain, behaviour and cognition. Nat Rev Neurosci. 2009;10(6):434–445. doi:10.1038/nrn2639

43. McEwen BS, Akil H. Revisiting the stress concept: implications for affective disorders. J Neurosci. 2020;40(1):12–21. doi:10.1523/JNEUROSCI.0733-19.2019

44. McEwen BS. Neurobiological and systemic effects of chronic stress. Chronic Stress (Thousand Oaks). 2017;1:2470547017692328. doi:10.1177/2470547017692328

45. Souza-Talarico JN, Marin MF, Sindi S, Lupien SJ. Effects of stress hormones on the brain and cognition: evidence from normal to pathological aging. Dement Neuropsychol. 2011;5(1):8–16. doi:10.1590/S1980-57642011DN05010003

46. Miller GE, Chen E, Parker KJ. Psychological stress in childhood and susceptibility to the chronic diseases of aging: moving toward a model of behavioral and biological mechanisms. Psychol Bull. 2011;137(6):959–997. doi:10.1037/a0024768

47. Calderón-Garcidueñas L, Leray E, Heydarpour P, Torres-Jardón R, Reis J. Air pollution, a rising environmental risk factor for cognition, neuroinflammation and neurodegeneration: the clinical impact on children and beyond. Rev Neurol (Paris). 2016;172(1):69–80. doi:10.1016/j.neurol.2015.10.008

48. Roveta F, Bonino L, Piella EM, Rainero I, Rubino E. Neuroinflammatory biomarkers in Alzheimer’s disease: from pathophysiology to clinical implications. Int J Mol Sci. 2024;25(22):11941. doi:10.3390/ijms252211941

49. Heneka MT, van der Flier WM, Jessen F, et al. Neuroinflammation in Alzheimer disease. Nat Rev Immunol. 2025;25(5):321–352. doi:10.1038/s41577-024-01162-8

50. Zotova E, Bharambe V, Cheaveau M, et al. Inflammatory components in human Alzheimer’s disease and after active amyloid-β42 immunization. Brain. 2013;136(9):2677–2696. doi:10.1093/brain/awt210

51. Boon BD, Hoozemans JJM, Lopuhaä B, et al. Neuroinflammation is increased in the parietal cortex of atypical Alzheimer’s disease. J Neuroinflammation. 2018;15(1):170. doi:10.1186/s12974-018-1222-6

52. Moonen S, Koper MJ, van Schoor E, et al. Pyroptosis in Alzheimer’s disease: cell type-specific activation in microglia, astrocytes and neurons. Acta Neuropathol. 2023;145(2):175–195. doi:10.1007/s00401-022-02518-7

53. Escartin C, Galea E, Lakatos A, et al. Reactive astrocyte nomenclature, definitions, and future directions. Nat Neurosci. 2021;24(3):312–325. doi:10.1038/s41593-020-00783-4

54. Han RT, Kim RD, Molofsky AV, Liddelow SA. Astrocyte-immune cell interactions in physiology and pathology. Immunity. 2021;54(2):211–224. doi:10.1016/j.immuni.2021.01.013

55. Wang L, Nykänen NP, Western D, et al. Proteo-genomics of soluble TREM2 in cerebrospinal fluid provides novel insights and identifies novel modulators for Alzheimer’s disease. Mol Neurodegener. 2024;19(1):1. doi:10.1186/s13024-023-00602-z

56. Kim JE, Knox M, Grill JD, et al. Virtual data collection strategies in research on Alzheimer’s disease and related dementias (ADRD). Innov Aging. 2025;9(5):igaf026. doi:10.1093/geroni/igaf026

57. Gershon RC, Wagster MV, Hendrie HC, Fox NA, Cook KF, Nowinski CJ. NIH toolbox for assessment of neurological and behavioral function. Neurology. 2013;80(11 suppl 3):S2–S6. doi:10.1212/WNL.0b013e3182872e5f

58. Toolbox Assessments, Inc. Cognition Assessments. NIH Toolbox. Accessed May 17, 2026. https://nihtoolbox.org/domain/cognition/

59. Juster RP, McEwen BS, Lupien SJ. Allostatic load biomarkers of chronic stress and impact on health and cognition. Neurosci Biobehav Rev. 2010;35(1):2–16. doi:10.1016/j.neubiorev.2009.10.002

60. Souza-Talarico JN, Perkhounkova Y, Hein M, Lee J, Hefti M, Sindi S. Allostatic load dynamics, Alzheimer’s disease biomarkers, and progression in individuals with mild cognitive impairment: findings from the Alzheimer’s Disease Neuroimaging Initiative. Alzheimers Dement (Amst). 2025;17(3):e70140. doi:10.1002/dad2.70140

61. Ho EH, Karpouzian-Rogers T, Ayturk E, Bedjeti K, Weintraub S, Gershon R. NIH Toolbox cognition performance in older adults with normal cognition, mild cognitive impairment, and mild dementia of the Alzheimer’s type: results from the ARMADA study. Arch Clin Neuropsychol. 2025;40(7):1279–1287. doi:10.1093/arclin/acaf045

62. Jutten RJ, Ho EH, Karpouzian-Rogers T, et al. Computerized cognitive testing to capture cognitive decline in Alzheimer’s disease: longitudinal findings from the ARMADA study. Alzheimers Dement (Amst). 2025;17(1):e70046. doi:10.1002/dad2.70046

63. Scott EP, Sorrell A, Benitez A. Psychometric properties of the NIH Toolbox Cognition Battery in healthy older adults: reliability, validity, and agreement with standard neuropsychological tests. J Int Neuropsychol Soc. 2019;25(8):857–867. doi:10.1017/S1355617719000472

64. Snitz BE, Tudorascu DL, Yu Z, et al. Associations between NIH Toolbox Cognition Battery and in vivo brain amyloid and tau pathology in non-demented older adults. Alzheimers Dement (Amst). 2020;12(1):e12018. doi:10.1002/dad2.12018

65. Casaletto KB, Umlauf A, Marquine M, et al. Demographically corrected normative standards for the Spanish language version of the NIH Toolbox Cognition Battery. J Int Neuropsychol Soc. 2016;22(3):364–374. doi:10.1017/S1355617716000041

66. Karr JE, Rodriguez JE, Goh PK, Martel MM, Rast P. The unity and diversity of executive functions: a network approach to life span development. Dev Psychol. 2022;58(4):751–768. doi:10.1037/dev0001324

67. Tourangeau R, Yan T. Sensitive questions in surveys. Psychol Bull. 2007;133(5):859–883. doi:10.1037/0033-2909.133.5.859

